# Hexamaps for Visualizing Age-Period-Cohort Data Trends

**DOI:** 10.1101/19011700

**Authors:** Hawre Jalal, Donald S. Burke

## Abstract

Age-period-cohort (APC) analyses often reveal important insights into patterns of disease incidence and mortality such as cancer. Both visual and analytical APC tools are available to reveal patterns by age, period and cohort. While developing new analytical methods of APC data is an active area of research, the choices of visual tools specific to APC data has been limited. In this study, we propose a “hexamap” as a new APC specific data visualization tool. This tool uses hexagons to address the unique challenge of APC data visualization, that is cohort= period - age. The approach is further illustrated using alcohol related mortality. Flexible Open-Source functions for implementation in R is also provided.

## Background

Age-period-cohort (APC) analyses is currently the mainstay to disentangle the effect of age, period and cohort on disease outcomes, such as cancer incidence and death. Because cohort = period – age, typical APC analysis involves an identification issue. Significant progress in addressing this issue has been achieved with many APC-specific analytical tools, such as ^1–6^ However, there is a lack of APC-specific data visualization tools. Most of APC data visualization rely on simple line charts or heatmaps on a regular grid that can only clearly present data on two of the APC axes.

In this paper, we propose hexagonal heatmaps as APC-specific data visualization tool that can overcome some of the challenges in current data display. We illustrate the hexagonal heatmaps with two examples. We also provide the R-code implementation of the tool to facilitate wider adoption and implementation.

### Hexagonal grids

Square grids are often used in Lexis diagrams to display APC data, where the data is arranged in a rectangular field and the rates or counts in each square is shown using different colors or shades (Figure 1A). [For simplicity, only an empty grid is shown here]. In this setup, age is often shown perpendicular to period while cohorts are being placed on the diagonal between age and period. Because of this uneven alignment, this display is less effective in revealing cohort patterns relative to age or period because (1) the square pixels only share a corner making it visually challenging to see the patterns along the diagonals relative to the pixels along the age and period axes that share a full side, and (2) this display creates a visual distortion where the spacing among cohorts seem to be wider than age or period. For example, if the data is arranged at one-year intervals, successive cohorts seem to be separated at 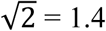 years, causing cohort patterns to appear diluted relative to those for age or period.

**Figure 1.**
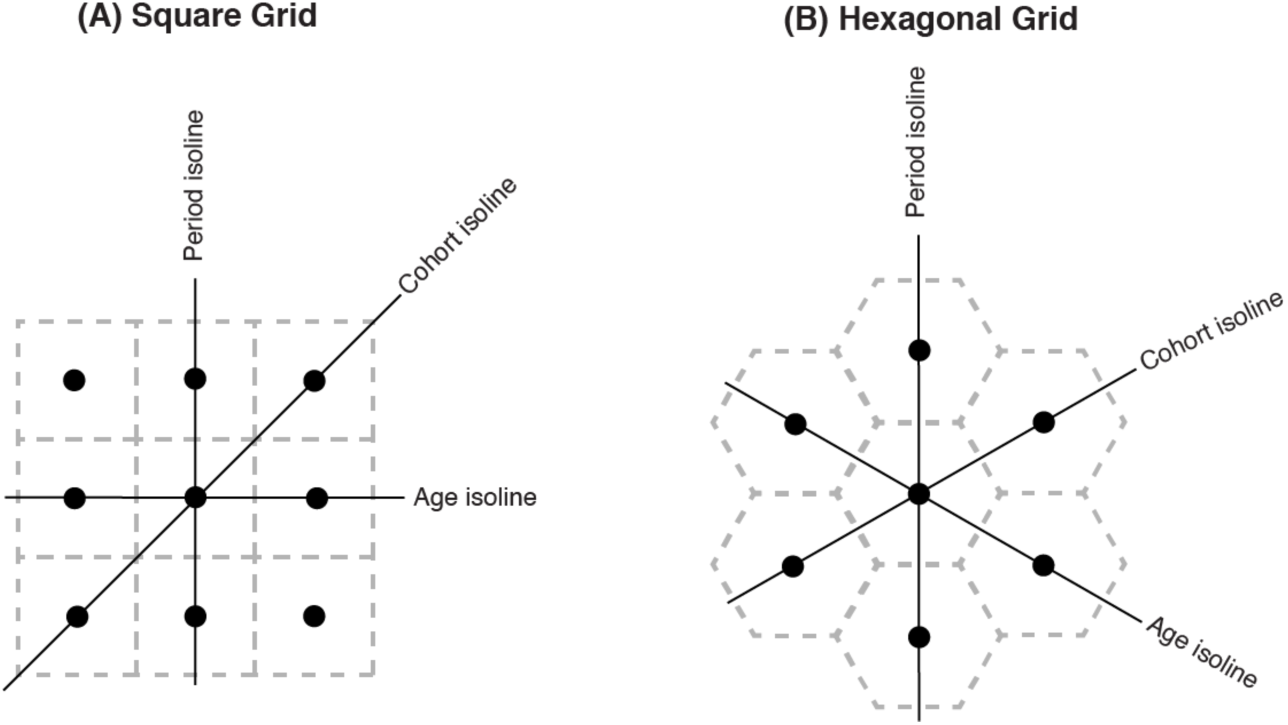
Age-period-cohort isoline orientation in a square vs. hexagonal data grid.

We propose a hexagonal grid (Figure 1B) to overcome the limitations of the traditional square grid. Hexagons are the most rounded shape that can tesselate (i.e., be repeated edge-to-edge without gaps to create an evenly spaced grid)^7^. Each cell in a hexagonal grid can have six neighbors. Thus, for each cell, one can easily navigate to how the patterns are changing with increasing or decreasing age, period or cohort. Specifically, because the hexagonal grid places the APC axes at equal angles from each other, the hexagonal pixels along any of the three dimensions share a side, facilitating visual pattern recognition along these axes. In addition, successive ages, periods and cohorts are equally spaced, thereby facilitating the visual display and interpretation of APC data along these axes.

### Plotting APC hexagons on the XY coordinate system

Using a hexagonal grid on a two dimensional medium or display requires computing the coordinates for the center of each hexagon on the Cartesian XY coordinate system. Here we follow the age, period and cohort isoline placements as shown in Figure 1 (B). We use these equations to compute the center for each hexagon based on age and period setting the distance between any two neighbouring hexagonal pixel centers to equal to one unit of time (e.g., one year):

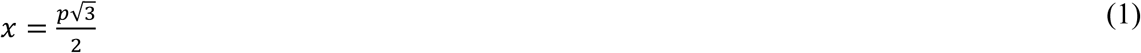

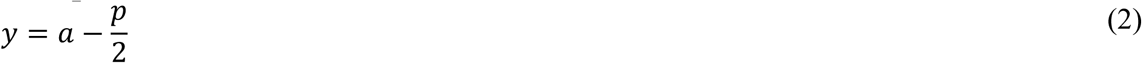

where *x* and *y* are the x and y coordinates on the Cartesian coordinate system, and *a* = age, and *p* = period. Because of our setup with period as vertical isolines, the x-coordinate of the hexagonal pixel is only a function of *p*, while the y-coordinate is a function of both *p* and *a* as they both determine the vertical position of the hexagonal pixels. Then, each hexagon can be plotted around its computed center using these XY coordinates for the hexagonal vertices (corners):

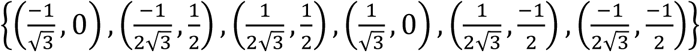

### Case study – alcohol related mortality in the United States from 1999 through 2017

To illustrate the Hexamap, we use alcohol-related mortality rates in the US from 1999 through 2017 as an example. The data was obtained from CDC-Wonder.^8^ Alcohol related poisoning is identified as alcohol overdose poisonings (International Classification of Disease 10^th^ Revision (ICD-10) codes of X45, X65 or Y15. The starting period is 1999, starting age is 0, and the interval is 1 year.

Figure 2 shows the hexamap as produced in R for alcohol-related mortality rates. Each data point at the intersection of age, period and cohort is represented as a hexagon. The APC isolines are placed at 5-year intervals. Alcohol related deaths are increasing among most age groups, especially after 2008. In 2017, the highest mortality rates were between ages 53-63. In addition to the increase of deaths by period, mortality rates seem especially higher among cohorts born after 1947.

**Figure 2:**
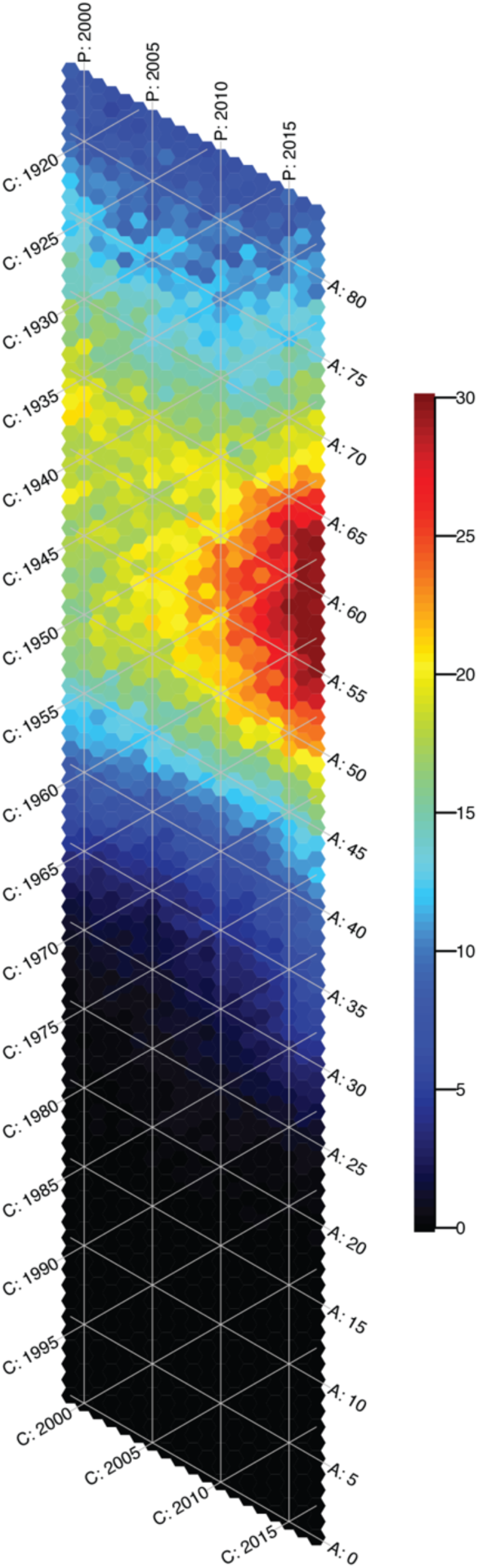
A hexamap showing the birth-year cohort patterns and the recent increase in alcohol-related mortality rates in the US from 1999 through 2017.

## Discussion

In this paper we propose a hexamap as a new way to visualize APC data. We illustrate how a hexagonal grid can overcome challenges of traditional visualization of APC data. We illustrate that using a hexagonal grid, one can compare patterns by changing age, period and cohort. We provide the functionality using R code in the supplement.

Researchers have often presented APC data using generic non-APC specific visualizations ranging from simple line charts to 3d plots ^9^. Line charts often show the incidence or mortality rates by age or period on the x-axis with individual lines presenting various cohorts. These line charts are challenging to interpret as they can only provide data readily on two of the three dimensions without a clear link between the remaining dimension. In addition, 3-d plots have been proposed where two of the dimensions are often age and period, and the third dimension is the incidence rate or mortality. The display of APC data in these visualizations is not necessarily clear given that one still needs to rotate the 3-d plot to 45 degrees to visualize the cohort patterns.

Hexagons are especially attractive for data visualization, because they are the most rounded object that creates a continuous grid. Hexagons has been recommended for visualizing data in many fields, such as ecological modeling and spatial analysis ^7^, showing bivariate statistical density distribution ^10^, and traffic pattern studies ^11^. In these fields, hexagons are often preferred because each data point can be compared readily to six neighboring data points compared to the four data points in traditional square grid presentation. In this paper, we illustrate how a hexagonal grid can be especially useful to visualize APC data because the data structure is naturally suited for a hexagonal.

It is important to note that our hexagonal grid places the APC axes at 60 degree angles. Thus, successive age, period and cohort isolines produce an equilateral triangular mesh. This equilateral mesh is equivalent to the Lexis’s equilateral diagram recommended in Lexis in 1875 12 and then by Weinkam and Sterling in 1991 ^13^ as a superior presentation of APC data to the standard square grid that produces right-angled triangular mesh. In addition, Weinkam and Sterling propose to overlay level curves that represent continuous regions with similar rate over the Lexis’ equilateral triangular mesh. However, this display is complicated as it involves multiple intersecting isolines of the various levels with the isolines of the APC axes. However, the use of the triangular mesh and the level-curves have been limited because of their complexity. Unlike the level-curves, the hexagons do not interfere with the isolines. Thus, we hope that researchers will find the hexamaps and the associated R code as a more practical and intuitive way to reveal patterns across the APC dimensions.

## Data Availability

The data was obtained from CDC-Wonder.

https://wonder.cdc.gov/

## Supplementary material R code

In this section we provide the R functions that can create hexamaps for any matrix of counts or rates by age and period. In summary, the hexamaps can be produced by running the `create_hexamap()` function which requires four parameters:

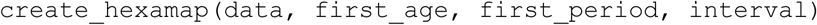

where `data` is a matrix containing the values to be plotted. The function expects the values to be arranged by age as rows and periods as columns. This pattern is consistent with other tools, such as the National Cancer Institute (NCI)’s online APC tool^14^ and the Surveillance, Epidemiology, and End Results (SEER) program that generally produces outputs in this structure.

Similarly, `first_age` is the first age group in the data, `first_period` is the first period in the data, and `interval` defines the interval that the data is spaced. The code requires the data to be equally spaced for both period and age. The `create_hexamap()` then creates the hexagons, colors them based on their relative values and then overlays the age, period and cohort isolines. Optionally, the user can modify how the starting values for the isolines, and how the isolines are spaced. In addition, the user can also modify some of the aesthetics, such as the color scales used to show the intensity of the values, the color and width of the lines, and the color and size of the fonts for the isolines. Other properties can also be modified by varying the underlying code which only requires base R.

For example, the code below reads the data and generates the hexamap for alcohol mortality rates in the US from 1979 through 2017 shown in Figure 2:

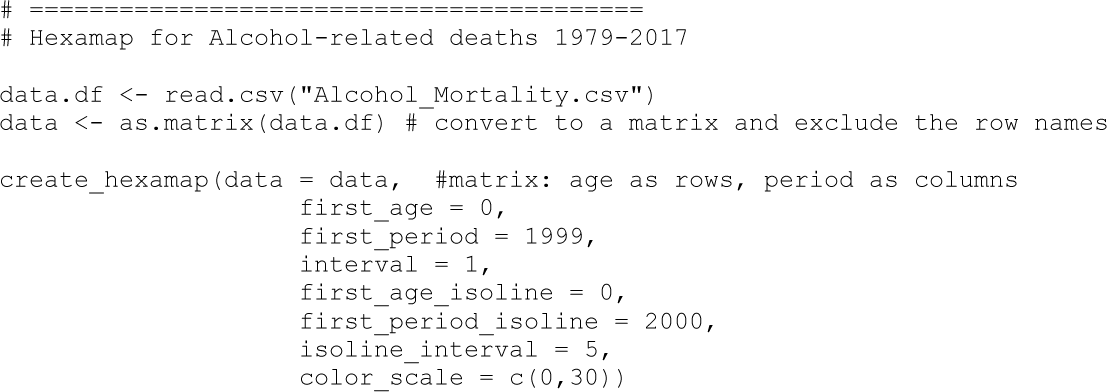

The code below shows the main hexamap function which is dependent on two functions to transform the X and Y coordinates to the hexagonal APC grid:

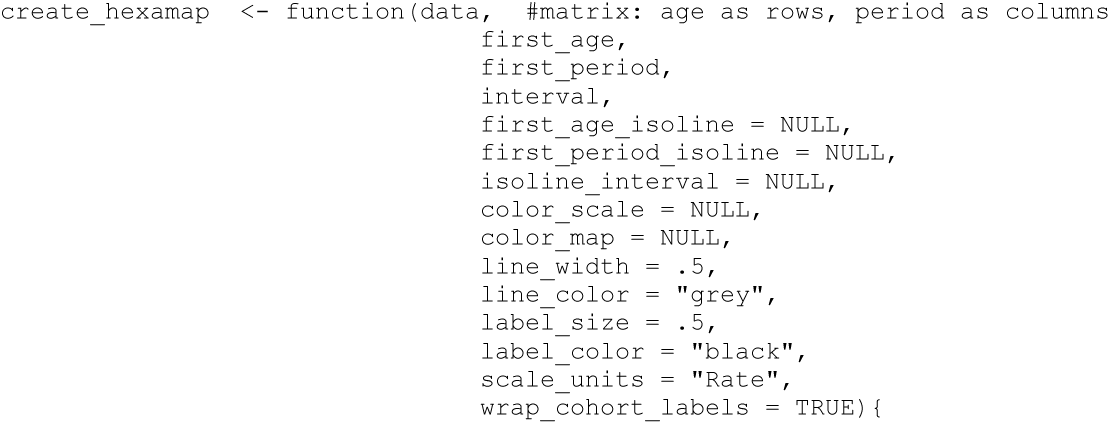

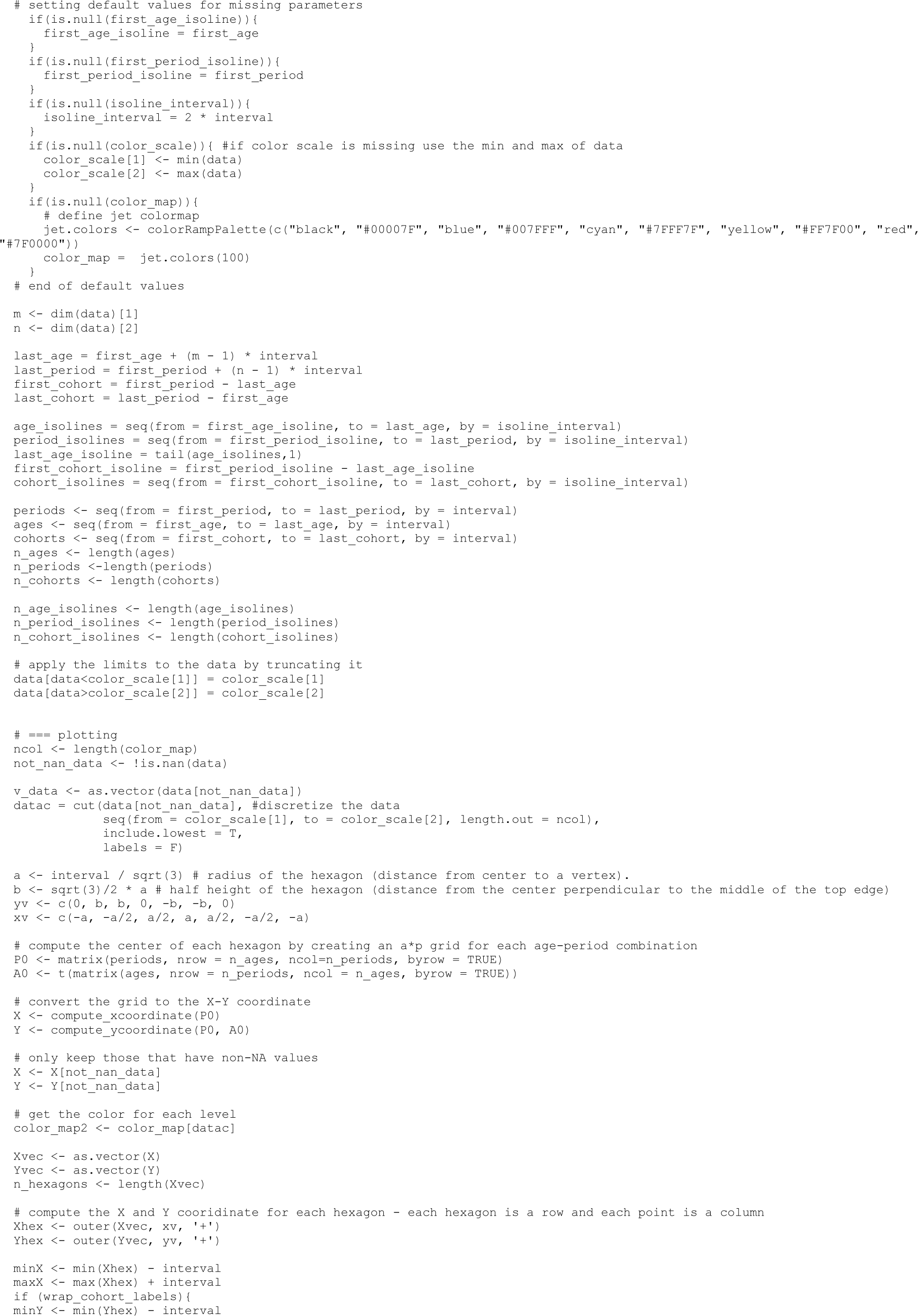

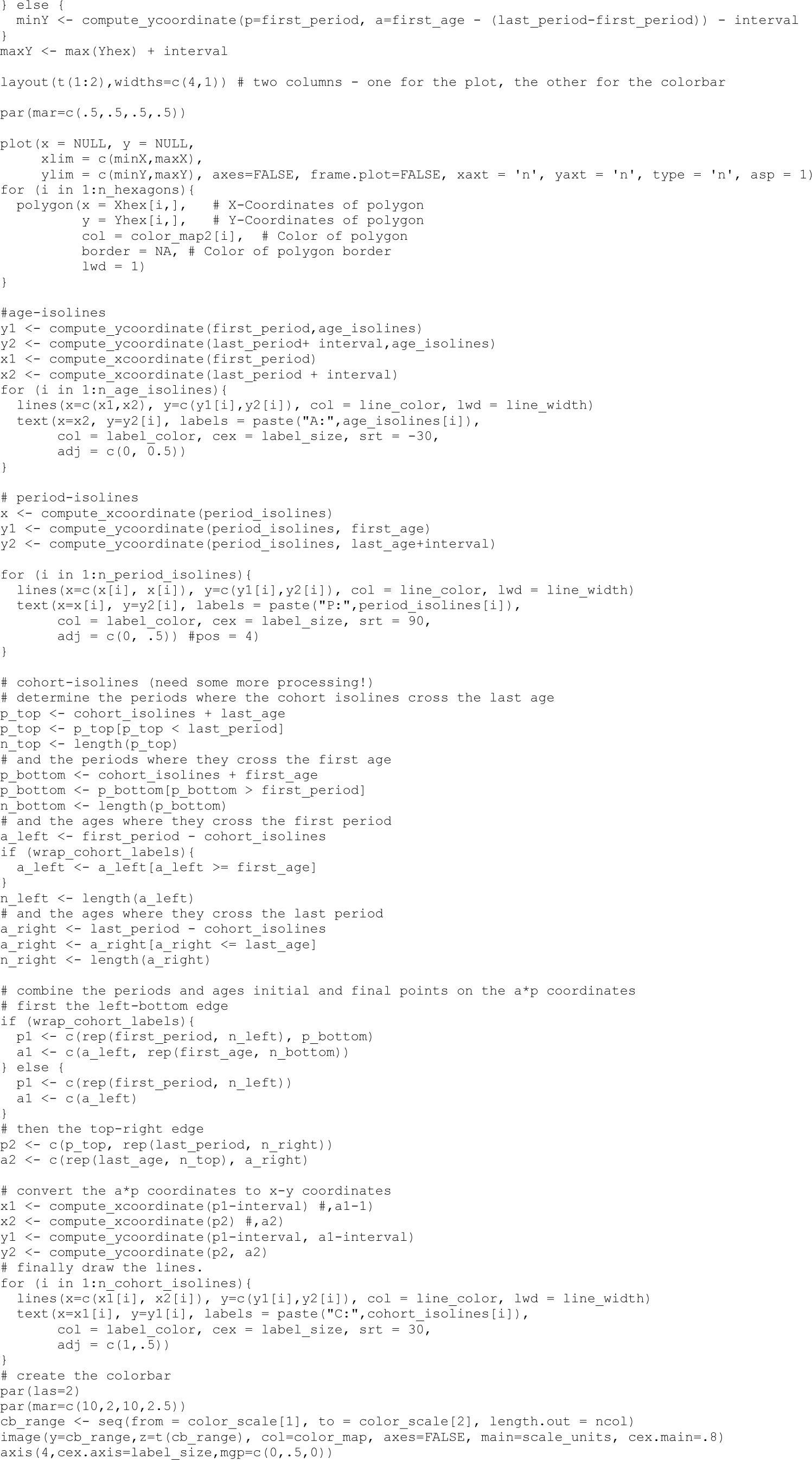

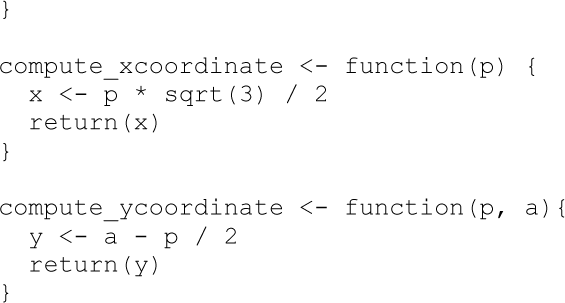

